# More Signal vs. More Noise - Comparing Full Text and Abstract as Inputs for Large Language Model-based Classification of Oncology Trial Eligibility Criteria

**DOI:** 10.64898/2026.03.10.26348010

**Authors:** Julia Weyrich, Fabio Dennstädt, Robert Förster, Christina Schröder, Daniel M. Aebersold, Daniel R. Zwahlen, Paul Windisch

**Author notes:** **Correspondence:** Julia Weyrich, Department of Radiation Oncology, Kantonsspital Winterthur, Brauerstrasse 15, Haus R, 8400 Winterthur.

## Abstract

**Purpose:** Large language models (LLMs) offer significant potential for automating the classification of clinical trials by eligibility criteria. However, a critical question remains regarding the optimal input data: while abstracts provide a condensed, high-density signal, full-text articles contain a much higher volume of information. It remains unclear whether the additional signal found in full texts improves classification performance or if the accompanying noise (in the form of thousands of words irrelevant to the question at hand in a complete manuscript) negatively affects the model’s reasoning capabilities.

**Methods:** GPT-5 was applied to classify 200 randomized controlled oncology trials from high-impact medical journals, labeling them whether patients with localized and/or metastatic disease were eligible for inclusion. Each trial was classified twice - once using only the abstract and once using the full text - and GPT-5’s outputs were compared with the ground-truth labels established by manual annotation. Performance was assessed by calculating and comparing accuracy, precision, recall, and F1 score, and the McNemar test was used to assess the statistical significance of the differences between the two input formats.

**Results:** For identifying trials including patients with localized disease, GPT-5 achieved an accuracy of 86% (95% CI: 81% - 91%; F1 = 0.90) when using abstracts and 92% (95% CI: 88% - 95%; F1 = 0.92) when using full texts (p = 0.027). Performance for detecting trials, which include patients with metastatic disease, was comparably high, with accuracies of 99% (95% CI: 99% - 100%; F1 = 1.00) based on abstracts and 98% (95% CI: 97% - 100%; F1 = 0.99) based on full texts. Overall accuracy for assigning combined labels per trial increased from 86% (95% CI: 81% - 91%) using abstracts to 92% (95% CI: 88% - 95%) using full texts (p = 0.027).

**Conclusion:** Providing full-text articles to GPT-5 significantly improved the classification of trial eligibility criteria. These findings suggest that, for this task, the benefit of the additional signal contained within the full text outweighed the potential for performance degradation caused by increased noise. Utilizing full-text analysis appears particularly valuable for extracting specific eligibility criteria in oncology that are frequently omitted or not explicitly described within the abstract.

## INTRODUCTION

Artificial intelligence (AI) is becoming increasingly important in oncology research and knowledge synthesis, particularly for extracting and analyzing complex, unstructured clinical data through automated text mining.^1, 2, 3^ Simultaneously, the exponential growth of biomedical publications makes it progressively difficult for clinicians and researchers to identify relevant studies or base clinical decisions on the totality of available evidence.^2^ Advances in natural language processing (NLP) offer the potential to automate the search for relevant information in clinical trials, thereby accelerating both literature reviews and evidence retrieval.^3, 4^

In oncology, NLP-based approaches can be utilized to extract specific inclusion and exclusion criteria that define a study population. One criterion of substantial clinical interest is disease stage - specifically, whether a trial enrolled patients with localized disease, metastatic disease, or both. Recent efforts to automate this classification have utilized various machine-learning methods and Large Language Models (LLMs). However, these studies have relied almost exclusively on abstracts, which are relatively short, structured, and cost-effective to analyze. ^5, 6, 7^

The reliance on abstracts presents a considerable limitation: critical eligibility details are often omitted or only partially described in the summary. While analyzing the full text of a publication could theoretically resolve this, the impact of providing significantly longer contexts to LLMs remains a subject of ongoing debate in the field. On one hand, the full text provides a much stronger signal, containing the explicit details necessary for accurate classification. On the other hand, a complete manuscript introduces a substantial amount of noise - thousands of words irrelevant to the specific clinical question - which may degrade model performance. This ambivalent character of noise has also been described in other domains of physiology and medicine, where it is often treated as an unwanted component and filtered out, for example in medical imaging, but it has also been shown to enhance the performance of certain biomedical systems.^8, 9^

The ability of LLMs to maintain reasoning performance while filtering through high volumes of irrelevant content is a critical factor in determining their utility for large-scale evidence synthesis. As the training of LLMs on datasets such as OpenWebText inevitably involves noisy data, it has been shown that LLMs are relatively robust to large amounts of random noise during training. However, this robustness is not directly linked to downstream performance.^10^

Consequently, it remains an open question whether the increased signal found in a full-text manuscript justifies the added computational cost and the potential for noise-induced errors in LLM-based workflows. This study investigates this trade-off by focusing on a subset of 200 oncological trials previously investigated in abstract-based analyses.^5, 6^ We applied a state-of-the-art LLM to these publications to classify whether patients with localized and/or metastatic disease were eligible for inclusion, comparing the performance of abstract-only inputs against full-text inputs.

Our hypothesis was that, for this specific task, the LLM would achieve higher classification accuracy when provided with the full text. While this has been demonstrated in a previous comparison of abstract-versus full text-based text mining for a non-LLM-based Named Entity Recognition (NER) system, we wanted to run this comparison for the modern text mining era, anticipating that the LLM’s capacity to locate the more detailed signal within the main text would outweigh the potential for performance degradation caused by the increased volume of irrelevant information..^11^

## METHODS

### Data selection

For this analysis, a previously annotated data set of 600 randomly-drawn oncology randomized controlled trial publications published between 2005 and 2023 across six major journals (*British Medical Journal, JAMA, JAMA Oncology, The Lancet, Lancet Oncology, New England Journal of Medicine*) was used as the foundation.^5^ From this data set, we drew a random sample of 200 trials excluding trials published in the Journal of Clinical Oncology, as the full texts of these publications were not available with the institutional licenses. No journal-level quotas were applied. The full text-PDFs of all 200 trial publications were downloaded from the respective journal websites, and their text was extracted using the PyMuPDF library. The corresponding code and dataset are available on GitHub.^12^

### Manual Annotation

To establish the ground truth, all papers had previously been manually annotated by a single author (P.W.) with the labels “LOCAL” and “METASTATIC”. Trials where patients with localized disease were eligible received the label “LOCAL” whereas trials allowing the inclusion of patients with metastatic disease received the label “METASTATIC”. Trials that included both localized or metastatic disease populations received both labels. Trials in which neither patients with localized nor metastatic disease were eligible (e.g. screening trials) received no label. Trials of tumor entities where the distinction between localized and metastatic diseases is not typically made (e.g., hematologic malignancies) were excluded.^5^ The previous annotation was based on the title and abstract. The full text was only to be consulted if title and abstract were deemed inconclusive to perform the classification. In a second step, another author (J.W.) independently annotated the 200 trials again, this time based on the full text. Inter-annotator agreement was calculated as the raw percent agreement, defined as the proportion of trials for which both annotators assigned identical label sets. Discrepancies were discussed between the two authors with a third author (D.R.Z.) acting as a judge in case of potential disagreements.

### LLM-based Classification

The performance of a commercially available OpenAI model (GPT-5, OpenAI, San Francisco, CA) with snapshot “gpt-5-2025-08-07”, temperature set to 1, and reasoning level set to high was then evaluated by having it label each trial as “LOCAL”, “METASTATIC”, both or none. The decision to use an OpenAI model was based on the prevalent use of these models at the time as well as the convenience of API access and lack of privacy concerns with the study data. For each trial, the abstract and full text were provided separately to the model, together with an instruction to output only the appropriate label(s) or none. The results were produced in a single batch run (without repetition) across all 200 trials. We refrained from performing multiple classification runs as a previous paper from the group had shown very consistent performance by LLMs for both classification and named-entity recognition tasks as long as the temperature was kept at or below 1.50.^13^

We used a zero-shot approach enforcing structured output. The system prompt was the following:

*“You will be provided with text excerpts from a cancer clinical trial. Your task is to classify patient eligibility based on disease stage. Specifically, determine whether the trial included:*

- *Patients with localized or locally advanced disease, i*.*e. cancer that has not formed distant metastases*
- *Patients with metastatic disease*

*Your output must be a valid Python-list, this list may contain 0, 1 or 2 elements. The allowed elements are:*

- *‘LOCAL’ = Patients with localized or locally advanced disease were eligible for the trial*
- *‘METASTATIC’ = Patients with metastatic disease were eligible for the trial If you are unsure, please provide your best guess*.

*Do NOT return any additional text, explanations or punctuation marks outside the list*.*”*

The user prompt was the respective text section.

To assess GPT-5’s accuracy in correctly predicting both labels (combined labels) right for a given trial, the list with the labels “LOCAL” and “METASTATIC” was condensed into the labels “Both” (LOCAL and METASTATIC), “Metastatic only” (METASTATIC), “Local only” (LOCAL) and “Neither”, when GPT-5 returned an empty list.

Accuracy, precision, recall and F1 score were calculated and compared for the labels separately as well as for the combined labels. P-values were derived for accuracy using the McNemar test, and 95% confidence intervals were computed for all metrics using normal approximation intervals.

The text extraction, the API calls and the statistical calculations were performed in Python (version 3.13.5) with the libraries pandas (version 2.3.1), pymupdf (version 1.26.4), numpy (version 2.3.1), openai (version 1.100.2), matplotlib (version 3.10.6), seaborn (version 0.13.2), and statsmodels (version 0.14.5). The complete code is available on GitHub.^12^

## RESULTS

### Inter-annotator agreement

The annotators agreed on 191 out of 200 trials (95.5%). Seven of the nine trials for which the annotations initially differed were changed from “METASTATIC” (P.W.) to both labels (J.W.), as only patients with metastatic disease were mentioned in the abstract, while the inclusion of patients with localized disease (e.g. “locally advanced unresectable”) was described in the full text.

### Distribution of trials

Figure 1a) shows the distribution of the 200 trials included in this analysis, categorized by whether patients with localized or metastatic disease were eligible for inclusion. Overall, 73.5% of the trials allowed the inclusion of patients with localized disease and 54% included patients with metastatic disease.

**Figure 1.**
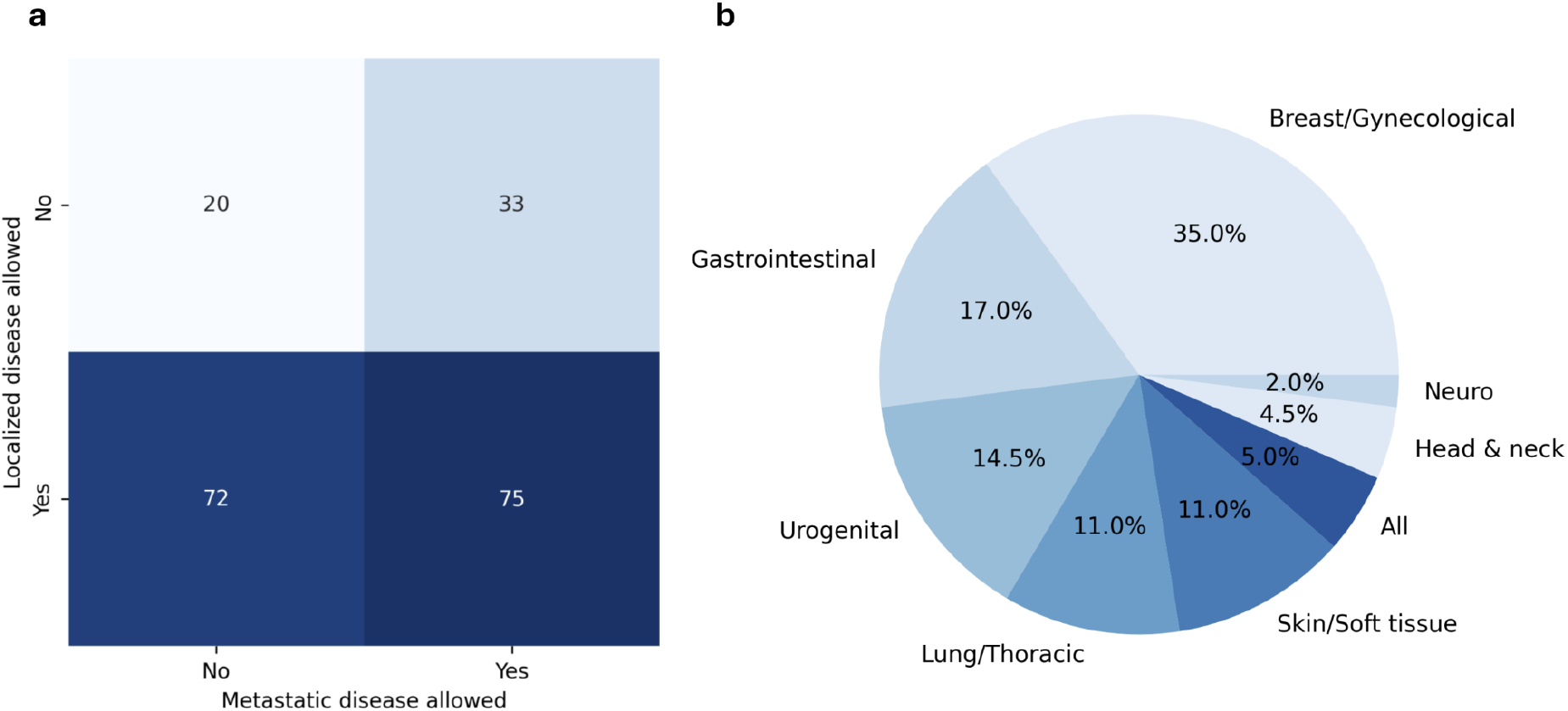
a) Distribution of the 200 trial publications, by whether they include patients with metastatic and /or localized tumors b) Distribution of grouped tumor entities in the dataset. Trials that allow for the inclusion of patients with any tumor entity e.g. trials for patients with painful bone metastases from any primary site are grouped under “All”.”

The distribution of grouped tumor entities (in percentage) is presented in Figure 1b. Breast and gynecological tumors accounted for the largest proportion of trials (35%), followed by gastrointestinal (17%) and urogenital (14.5%) tumors.

### Classification Performance

When provided with the abstracts of the trial publications, GPT-5 achieved an accuracy of 0.86 and an F1 score of 0.90 for predicting whether a trial included patients with localized disease (precision: 0.98, recall: 0.83). For metastatic disease, the accuracy was 0.99 and the F1 score was 1.00 (precision: 0.99, recall: 1.00)

When analyzing the full texts, GPT-5 achieved an accuracy of 0.92 and an F1 score of 0.94 in predicting the inclusion of patients with localized disease (precision: 0.96, recall: 0.92). For metastatic disease the accuracy was 0.98 and the F1 score 0.99 (precision: 0.97, recall: 1.00)

The accuracy of the prediction whether patients with localized disease were included based on full text was significantly higher than based on the abstract (p = 0.0266). The difference in accuracy, when predicting whether patients with metastatic disease were eligible was not significant (p = 0.5)

All performance metrics, including 95% confidence intervals, are presented in Table 1. The corresponding confusion matrices are shown in Figure 2.

**Table 1:** Performance Metrics of GPT-5 based on the Abstract and based on the full texts. Abbreviation: 95% CI = 95% Confidence Interval.

| Table 1 | Accuracy (95% CI) | Precision (95% CI) | Recall (95% CI) | F1 Score (95%) |
| --- | --- | --- | --- | --- |
| **Based on Abstract** |  |  |  |  |
| Localized disease | 0.86 (95% CI: 0.81 - 0.91) | 0.98 (95% CI: 0.95 - 1.00) | 0.83 (95% CI: 0.77 - 0.89) | 0.90 (95% CI: 0.85 - 0.94) |
| Metastatic disease | 0.99 (95% CI: 0.99 - 1.00) | 0.99 (95% CI: 0.97 - 1.00) | 1.00 (95% CI: 1.00 - 1.00) | 1.00 (95% CI: 0.99 - 1.00) |
| **Based on full text** |  |  |  |  |
| Localized disease | 0.92 (95% CI: 0.88 - 0.95) | 0.96 (95% CI: 0.93 - 1.00) | 0.92 (95% CI: 0.87 - 0.96) | 0.94 (95% CI: 0.91 - 0.97) |
| Metastatic disease | 0.98 (95% CI: 0.97 - 1.00) | 0.97 (95% CI: 0.94 - 1.00) | 1.00 (95% CI: 1.00 - 1.00) | 0.99 (95% CI: 0.97 - 1.00) |

**Figure 2.**
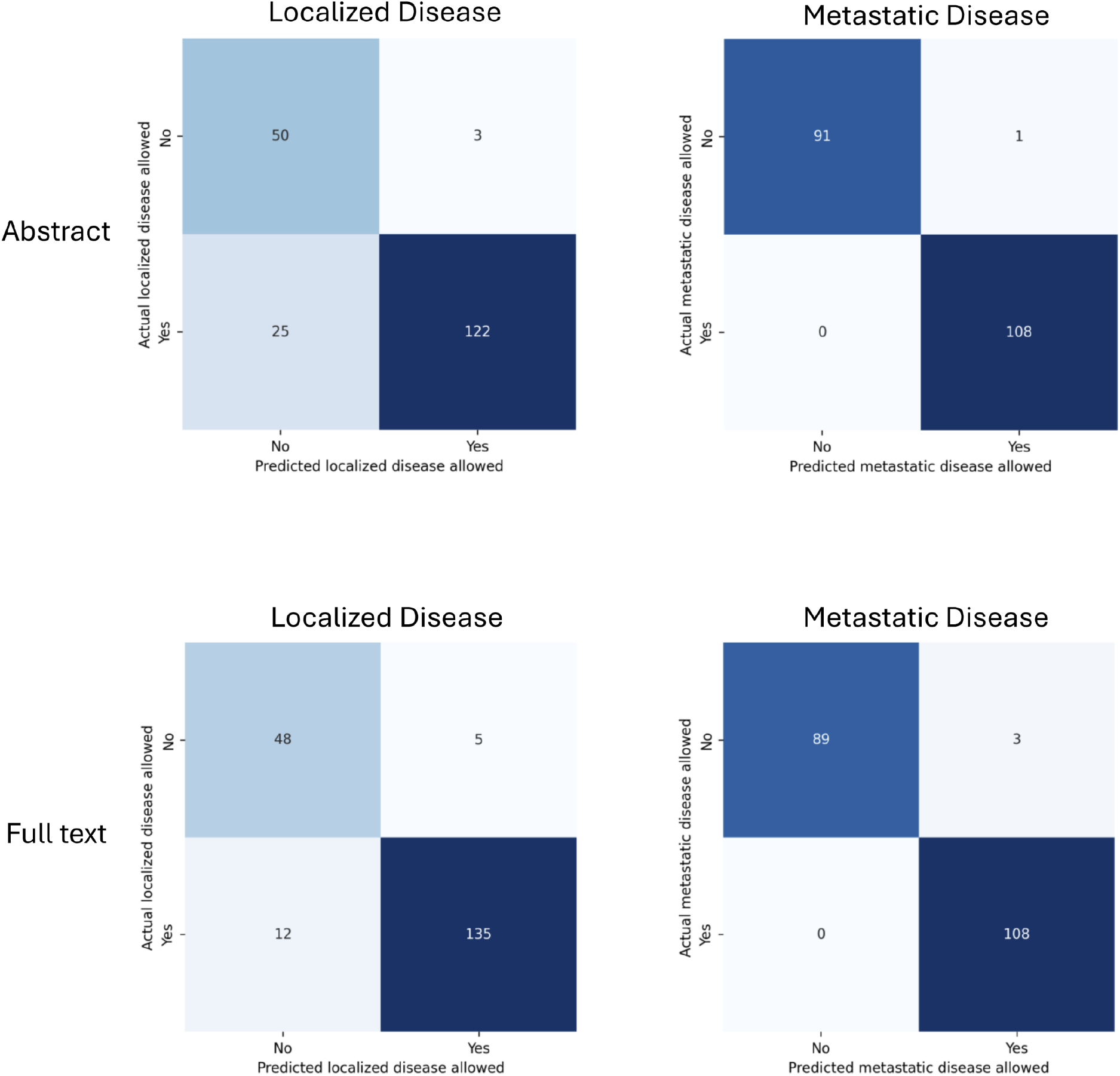
Confusion matrices showing the performance of GPT-5 when using abstracts (above) versus full texts (below) to predict trial eligibility for patients with localized disease (left) and metastatic disease (right). The corresponding performance metrics are presented in Table 1.

Regarding the combined labels, GPT-5 achieved an overall accuracy of 86% when provided with the abstracts and 92% when provided with the full texts of the publications, with the difference being statistically significant (p = 0.027).

Figure 3 displays the confusion matrices for GPT-5 when using abstracts versus full texts for assigning all labels to a trial, the corresponding performance metrics are presented in Table 2.

**Table 2:** Performance Metrics of GPT-5 when assigning all labels combined. Abbreviation: 95% CI = 95% Confidence Interval.

| Table 2 | Accuracy (95% CI) | Precision (95% CI) | Recall (95% CI) | F1 Score (95%) |
| --- | --- | --- | --- | --- |
| **Based on Abstract** |  |  |  |  |
|  | 0.86 (95% CI: 0.81 - 0.91) |  |  |  |
| Both |  | 0.98 (95% CI: 0.94 - 1.02) | 0.67 (95% CI: 0.56 - 0.77) | 0.79 (95% CI: 0.74 - 0.85) |
| Local only |  | 0.97 (95% CI: 0.94 - 1.00) | 1.00 (95% CI: 1.00 - 1.00) | 0.99 (95% CI: 0.97 - 1.00) |
| Metastatic only |  | 0.57 (95% CI: 0.44 - 0.70) | 1.00 (95% CI: 1.00 - 1.00) | 0.73 (95% CI: 0.66 - 0.79) |
| Neither |  | 1.00 (95% CI: 1.00 - 1.00) | 0.85 (95% CI: 0.69 - 1.00) | 0.92 (95% CI: 0.88 - 0.96) |
| **Based on full text** |  |  |  |  |
|  | 0.92 (95% CI: 0.88 - 0.95) |  |  |  |
| Both |  | 0.94 (95% CI: 0.88 - 1.00) | 0.84 (95% CI: 0.76 - 0.92) | 0.89 (95% CI: 0.84 - 0.93) |
| Local only |  | 0.99 (95% CI: 0.96 - 1.00) | 1.00 (95% CI: 1.00 - 1.00) | 0.99 (95% CI: 0.98 - 1.00) |
| Metastatic only |  | 0.73 (95% CI: 0.60 - 0.86) | 0.97 (95% CI: 0.91 - 1.03) | 0.83 (95% CI: 0.78 - 0.88) |
| Neither |  | 1.00 (95% CI: 1.00 - 1.00) | 0.80 (95% CI: 0.62 - 0.98) | 0.89 (95% CI: 0.85 - 0.93) |

**Figure 3.**
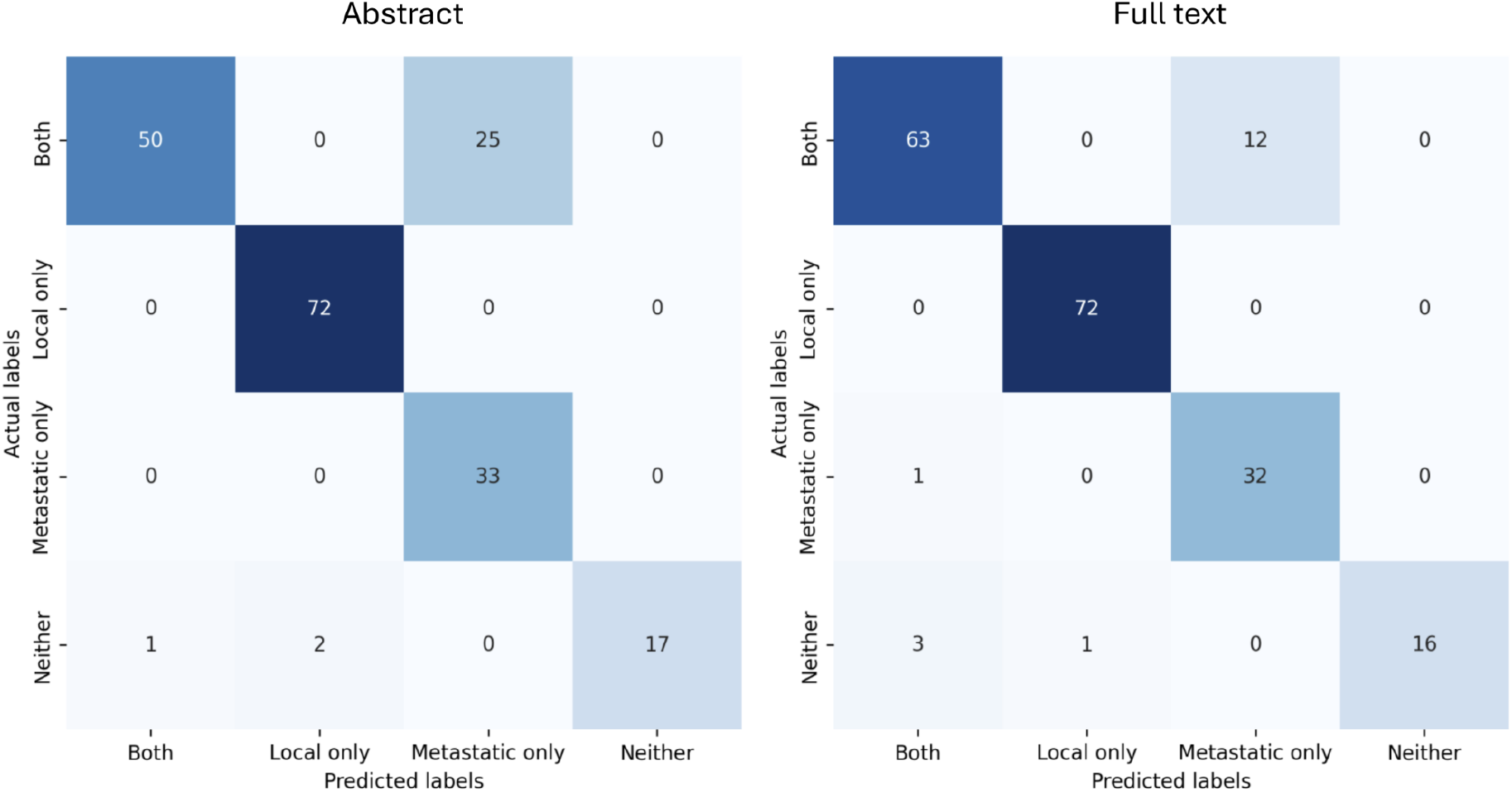
Confusion matrices showing the performance of GPT-5 when using abstracts (left) versus full texts (right) to predict the combined labels of patient eligibility. The corresponding performance metrics are presented in Table 2.

## DISCUSSION

When provided with the full texts rather than the abstracts alone, GPT-5 achieved a significantly higher accuracy in predicting whether patients with localized disease were included across the 200 oncological trial publications in our dataset (0.92 vs. 0.86, p = 0.027).

In predicting the inclusion of patients with metastatic disease, GPT-5 performed nearly on par with human annotation. Interestingly, its precision was slightly higher when classifications were based on the abstracts only (0.99) than on full texts (0.97), although this difference was not statistically significant (p=0.50).

The few trials (n=3) that GPT-5 misclassified as including patients with metastatic disease based on the full text, had been labeled as “neither” in the manual annotation (for example, screening trials or postoperative trials after tumor resection)

Considering the combined labels (“all labels”, Figure 3) most misclassifications involved trials that enrolled both patients with localized and metastatic disease. In these cases, GPT-5 often predicted only the metastatic label. Providing GPT-5 with the full text reduced this type of misclassification by half (6%, n=12) compared to when using the abstract alone (13.5%, n=25). This difference was statistically significant (p=0.027)

The identical p-values for the localized-only and combined labels arise because trials misclassified as including patients with metastatic disease were also misclassified as including patients with localized disease (“Neither” classified as “Both”). This means that the metastatic component did not affect the accuracy or the p-value of the combined-label analysis.

The improved accuracy of GPT-5’s prediction when provided the full texts, compared with the abstracts, likely reflects abbreviated reporting of study populations in abstracts. In trials enrolling both patients with localized and metastatic disease, abstracts frequently emphasize the metastatic population, while the inclusion of patients with localized disease (that is usually locally advanced and/or unresectable) is often only described in the main text. This pattern was also observed during the manual annotation process and accounted for most of the disagreements between the two annotators, as P.W. relied primarily on titles and abstracts, while J.W. used the full texts. In cases of disagreement, the full-text-based annotation or protocol was used as the ground truth to ensure that the labels reflect the most complete available information.

Overall, the gain in recall when GPT-5 was provided with the full texts outweighs the small loss in precision compared with abstracts-only classification. Consequently, GPT-5 reached 92% accuracy for predicting the combined labels based on the full texts versus 86% accuracy when using the abstracts alone.

Previous studies have classified oncological trials according to whether patients with localized or metastatic disease were eligible, using machine-learning approaches and different LLMs. However, these studies relied exclusively on abstracts, as abstracts are more readily available and less resource-intensive to process.^6^ Notably, GPT-5 already achieves higher accuracy on abstract-based classification than many earlier models.^6^ However, it remains unclear whether other LLMs would similarly benefit from full-text inputs in this classification task compared to abstract-only inputs. Models with comparable or greater robustness to noisy input might therefore achieve a similar or potentially higher performance than GPT-5 when provided with the full-text publications. Our results indicate that GPT-5’s performance is robust enough to filter through the noise of a complete manuscript to locate specific criteria. This suggests that the common reliance on abstracts in NLP research, while cost-effective, may lead to an underestimation of an LLM’s true performance potential in evidence synthesis.

This analysis has several limitations. First, it is limited to 200 oncological randomized controlled trials published in five journals that require structured abstracts; therefore, results may not be generalizable to other trial types or to articles without structured abstracts. However, structured abstracts are the standard in high-impact medical literature, making these results relevant to the majority of publications.

Second, because the model’s training cutoff postdates the publication of the included trials, we cannot rule out the influence of data leakage, where the model may have encountered these specific studies during its pre-training phase. However, this effect would likely have reduced the performance gap between abstract and full-text inputs, suggesting that the differences reported here represent a conservative estimate of the true benefit of full-text analysis.

Third, we did not assess GPT-5’s stability across repeated runs or different prompts; all classifications were generated in a single run, and performance may differ for simpler models with smaller context windows. Nonetheless, previous research indicates that high-performance LLMs demonstrate high consistency in structured classification tasks at low temperatures, and the use of a state-of-the-art model serves as a benchmark for the current technological upper bound.

Fourth, trials published in the Journal of Clinical Oncology had to be excluded due to institutional paywalls. While this reduces the journal diversity of our sample, the included journals represent a broad cross-section of top-tier medical research, and there is no reason to assume that the reporting style in JCO would fundamentally alter the “signal vs. noise” trade-off observed here.

Fifth, the full-text publications were converted into plain text before being provided to GPT-5. As a result, certain information - such as images, figures, and other non-textual elements - was not available to the model. Future studies could address this limitation by using vision-language models that allow the direct processing of the publications as PDF documents, thereby including both textual and visual information into the analysis.

Our findings indicate that using the full text in text mining is reasonable, especially when the information of interest is reported inconsistently or incompletely in abstracts. Whether this approach is worth it for a given task depends on the nature of the task and the information. Classifying a trial’s primary endpoint may work equally well - or even better - when based solely on the abstract, as primary endpoints are typically reported in abstracts. Conversely, features that are rarely mentioned in abstracts but consistently described in the full texts would probably benefit more from full-text classification.

Allowing GPT-5 to express uncertainty or output an “unclassifiable” label could be explored to investigate whether restricting it to higher-confidence predictions improves accuracy and reduces hallucinations. However, a recent study by Huang et al. suggests that the uncertainty estimation alone does not reliably indicate the correctness of a LLM’s output and proposes the incorporation of behavioral testing as a more promising approach.^14^

It would also be interesting to extend the classification to additional, more granular criteria - for example, primary endpoint, type of intervention, or different eligibility criteria - to determine which features benefit most from full-text input.

In conclusion, using full-text articles significantly improved classification accuracy compared to abstracts alone. For the task of identifying trial eligibility, the increased volume of information in the full text provides a critical signal that justifies the added computational context.

## Ethics approval and consent to participate

Not applicable

## Availability of data and materials

All data and code used to obtain this study’s results have been uploaded to https://github.com/JuliaWy/Full-text-vs-Abstract.

## Author contributions

Conceptualization, P.W., F.D.; methodology, J.W., P.W.; formal analysis, J.W.; data curation, J.W.; writing—original draft preparation, J.W., P.W.; writing—review and editing, F.D., R.F., C.S., D.M.A., D.R.Z.; supervision, D.R.Z..; project administration, D.R.Z.;

All authors read and approved the final manuscript

